# Cellular and humoral Immune response to mRNA COVID-19 vaccination in subjects with chronic lymphocytic leukemia

**DOI:** 10.1101/2021.11.04.21265948

**Authors:** Zoe L. Lyski, Myung S. Kim, David Xthona Lee, Hans-Peter Raué, Vikram Raghunathan, Janet Griffin, Debbie Ryan, Amanda E. Brunton, Marcel E. Curlin, Mark K. Slifka, William B. Messer, Stephen E. Spurgeon

**Affiliations:** Department of Molecular Microbiology & Immunology, Oregon Health & Science University (OHSU), Portland, OR 97239, USA; Knight Cancer Institute, Oregon Health & Science University (OHSU), Portland, OR 97239, USA; Division of Neuroscience, Oregon National Primate Research Center, Oregon Health & Science University, Beaverton, Oregon, USA; OHSU-PSU School of Public Health, Portland, OR 97239, USA; Department of Medicine, Division of Infectious Diseases, Oregon Health & Science University (OHSU), Portland, OR 97239, USA

## Abstract

Chronic Lymphocytic Leukemia (CLL) is predominantly a B-lymphocyte leukemia associated with immune defects that are often exacerbated by CLL directed therapies. SARS-CoV-2 infection poses a significant risk of illness or mortality to CLL patients, and while SARS-CoV-2 vaccines are highly effective in immunocompetent individuals, efficacy varies substantially in immunocompromised patients, including those with CLL. To date, studies of COVID-19 vaccine immune responses in immunocompromised hosts have largely relied on semi-quantitative antibody titers that only partially characterize vaccine-elicited immune responses and do not measure B or T-cell specific responses that may also play a protective role in vaccinees. Here, we report RBD-specific antibody as well as B-cell and T-cell responses in an observational cohort of sixteen CLL subjects who received mRNA vaccination against SARS-CoV-2, finding a strong association between CLL treatment and vaccine immunogenicity, with important implications for vaccination timing in the context of CLL treatment or recovery from prior treatment.

## Introduction

Severe acute respiratory syndrome coronavirus 2 (SARS-CoV-2), the causative agent of COVID-19 is of special concern to patients with hematological malignancies such as chronic lymphocytic leukemia (CLL). CLL is predominantly a B lymphocyte leukemia that over time impairs normal B-cell function and antibody production, leaving patients at increased risk of prolonged severe infections or death. Prolonged infection is additionally problematic because viruses may persist in patients for long periods of time in the presence of sub-neutralizing antibodies, possibly contributing to the development of viral variants of concern that can partially escape antibody-mediated immunity.^1-3^ Many patients with CLL not only suffer immune dysregulation from the disease, but their immune systems are further disrupted by the effects of CLL specific treatments, many of which further attenuate immune responses. There are now three vaccines for SARS-CoV-2 approved in the United States^4^, with high immunogenicity observed in immunocompetent subjects, leading to immunity that is associated with lower rates of severe infection.^5-7^ However, the immunogenicity and efficacy of these vaccines in immunocompromised hosts is less well characterized overall, and because immunocompromise is very heterogeneous, disease specific evaluation of vaccine immunogenicity and efficacy is imperative. The effectiveness of vaccines in immunocompromised patients, not included in vaccine trials, is critical in preparing the health care system for potential challenges and guiding individual patient decision during ongoing waves of SARS-CoV-2 as well as future emerging pathogens.

The post-immunization dynamics in CLL patients is different than that observed in healthy subjects. Attenuated humoral responses to vaccination have been previously documented for pneumococcal conjugated vaccine (PCV13), pneumococcal polysaccharide vaccine (PPSV23), and influenza A and B vaccination.^8-10^ There are several studies to date documenting impaired vaccine response to SARS-CoV-2 vaccines in patients with hematologic malignancies based on semi-quantitative antibody levels. Notably, patients with CLL have among the lowest immune responses, that are influenced by disease status, immunoglobulin (IgG) levels and active or recent therapies.^11-14^ In particular, treatment with anti-CD20 directed monoclonal antibodies is associated with very poor vaccine response. Additional treatment regimens incorporating Bruton tyrosine kinase inhibitors (BTKi) also affect vaccine response, a finding observed in the context of other vaccines as well.^15^

Here, we interrogated the cellular and humoral immune response to novel vaccine antigen, BNT162b2 (Pfizer-BioNTech) or mRNA-1273 (Moderna) in CLL subjects. In response to vaccination, immunocompetent individuals generate an antigen-specific response that results in cellular and humoral memory that persists long after vaccination^16^ including CD4+ T-cells, CD8+ T-cells, and two distinct long-lived populations of B-cells. The first, long-lived plasma cells (LLPC), traffic to the bone marrow and continuously secrete the antibodies that comprise polyclonal immune serum. The second B-cell population elicited by vaccination or natural infection are memory B-cells (MBCs). These cells do not secrete antibodies, but instead circulate in peripheral blood and traffic through lymph nodes surveying for invading pathogens. If they encounter a recognized pathogen, they respond by rapidly proliferating into antibody secreting cells. MBCs are especially important in the face of waning antibody titers or the emergence of new variants that might escape neutralization by serum antibodies because they are poised to rapidly respond with a new population of antibodies in the face of new infection^17^

In this longitudinal cohort study, cellular and humoral immune response to Pfizer-BioNTech or Moderna was evaluated in 16 subjects with CLL to better understand the extent of immune dysfunction caused by CLL and treatments. As a comparator, humoral recall response to the childhood antigen, measles was also evaluated. Subjects 18 years and older, with CLL and without a known history of COVID-19 infection, were enrolled at a single clinical site prior to receiving, the Moderna or Pfizer-BioNTech 2-dose SARS-CoV-2 mRNA vaccine series. This study reports the presence and magnitude of humoral and cellular immune responses including quantitative receptor-binding domain (RBD)-specific antibody titers, RBD-specific MBC frequency following *in vitro* stimulation and functional, TNF-α and INF-γ secreting spike peptide-specific CD4+ and CD8 T-cells at baseline (prior to vaccination) and for the majority of subjects around one-month (24-103 days) following two-dose mRNA vaccination series and compares the response. In an effort to better understand factors associated with vaccine response, baseline clinical and laboratory characteristics were assessed.

## Results

### Study participants

All 16 participants enrolled in the study received the first dose of either the BNT162b2 mRNA vaccine (Pfizer-BioNTech) or the mRNA-1273 vaccine (Moderna) between January 2, 2021 and April 13, 2021 and received their second dose 21-40 days later. Eight of 16 participants (50%) were undergoing CLL treatment, 4 of 16 (25%) patients were CLL treatment naïve and 25% (4 of 16) subjects were being observed after prior treatment (Table 1). Of the patients on active treatment, 6 of 8 (75%) were on BTKi monotherapy while the remaining 2 patients were on anti-CD20 monoclonal antibodies (mAb) combined with BCL2 inhibitor and BTKi respectively. Of the patients under observation, 3 of 4 patients (75%) had received treatment within the past 12 months. Eleven patients had previously received anti-CD20 mAb with 9 patients receiving a last dose more than 12 months prior to vaccination and 2 patients receiving the last dose 6 to 12 months prior to vaccination. Baseline serum IgG level was 466 mg/dL (median, IQR 85-918) and baseline absolute lymphocyte count was 2.91 K/cu mm (median, IQR 0.33-87). Individual results of baseline labs and summary of vaccine response are shown in Table 2.

**Table 1.** Summary subject demographic table. Summary data for 16 CLL subjects included in this study at the time of enrollment.

|  |  |
| --- | --- |
|  | N= 16<br>(median, IQR) |
| **Age** | 64.5 (60-75) |
| **Sex** |  |
| Male | 10 |
| Female | 6 |
| **Race** |  |
| White | 15 |
| Asian/White | 1 |
| **Treatment status** |  |
| Treatment naïve | 4 |
| Active treatment | 8 |
| Observation after prior treatment | 4 |
| **Previous lines of treatment (n=12)** | 1 (0-4) |
| **Current treatment (n=8)** |  |
| BCL2 inhibitor + anti-CD20 | 1 |
| BCL2 inhibitor + BTK inhibitor | 1 |
| BTK inhibitor | 6 |
| **Time since previous anti-CD20 (n=11)** |  |
| 6-12 months | 2 |
| >12 months | 9 |
| **Previous IVIG (n=6)** |  |
| ≤ 6 months | 0 |
| >6 months | 6 |
| **IGHV (n=8)** |  |
| Mutated | 5 |
| Unmutated | 3 |
| **Baseline IgG (n=10)** | 466 (85-918) |
| **Baseline ALC (n=14)** | 2.91 (0.33-87) |
| **Rai stage at diagnosis** |  |
| 0 | 9 |
| I | 5 |
| II | 1 |
| III | 0 |
| IV | 1 |

**Table 2.** Summary table of subject immune response to mRNA COVID-19 vaccination as well as recall response to measles antigen. An + indicates a response above the limit of detection, a - indicates a response below the limit of detection. For T-cell specific responses, a + indicates an increase in spike specific T-cells compared to baseline, a – indicates no change (or decrease) in spike specific T-cells following vaccination. Current treatment status, CD20 Ab treatment as well as clinical values taken at baseline (time of enrollment) when available. A dash for clinical values indicates that baseline values are not available. For clinical values, the normal range is indicated. N: Treatment Naïve, C: Currently on treatment, O (6): Observation, last treatment within 6 months, O (6-12): Observation, 6-12 months since last treatment, O (>12): Observation, more than 12 months since last treatment.

SARS-CoV-2 Measles
| ID | Age Range | Vaccine | Ab | CD4 | CD8 | MBC | Ab | MBC | Treatment | CD20 Ab | ALC<br>1-4.8 | IgG<br>700 - 1600 mg/dL | CD19+<br>4-17 (%) | IgD-CD27+<br>5-21 (%) | CD4<br>30-60 (%) | CD8 (%)<br>10-30% |
| --- | --- | --- | --- | --- | --- | --- | --- | --- | --- | --- | --- | --- | --- | --- | --- | --- |
| 1 | 60 - 64 | P | - | - | - | - | - | - | N | N | 48 | 85 | 96 | 3.6 | 2 | 0.7 |
| 2 | 60 - 64 | P | - | - | + | - | + | + | N | N | 21 | - | 76 | 59 | 9.3 | 10 |
| 3 | 45 - 50 | ? | + | + | - | - | - | - | C | Y ($\leq 12$) | 1 | - | 2.1 | 17 | 39 | 33 |
| 4 | 75-80 | M | - | - | - | - | + | - | N | N | 87 | 573 | - | - | - | - |
| 5 | 80-85 | P | - | - | + | - | + | + | C | Y ($> 12$) | 5.8 | - | - | - | 19 | 3.8 |
| 6 | 65-69 | M | - | + | + | - | + | + | C | N | 2.9 | 918 | 30 | 2.3 | 41 | 19 |
| 7 | 60-64 | P | - | + | + | - | + | - | O (6) | Y ($> 12$) | 1.6 | - | 0.004 | 0 | 86 | 8 |
| 8 | 65-69 | P | - | - | - | - | + | - | C | Y ($> 12$) | 0.43 | 526 | 3 | 22 | 35 | 47 |
| 9 | 65-69 | ? | - | + | + | - | + | - | C | Y ($\leq 12$) | 1.3 | - | 0.069 | 0 | 62 | 21 |
| 10 | 60-64 | P | - | + | - | - | - | - | C | Y ($> 12$) | 30 | 262 | 65 | 2.4 | 23 | 8 |
| 11 | 60-64 | P | + | + | + | - | + | + | N | N | 17 | 780 | 83 | 0.21 | 13 | 2.1 |
| 12 | 60-64 | P | + | + | + | + | + | - | O (6-12) | Y ($> 12$) | 0.75 | 593 | 8 | 3.8 | 42 | 15 |
| 13 | 70-74 | P | - | + | - | - | + | - | C | Y ($> 12$) | 5.8 | 101 | 45 | 1.5 | 20 | 20 |
| 14 | 65-69 | P | - | + | - | - | + | - | O ($> 12$) | Y ($> 12$) | 0.33 | 405 | - | - | 77 | 9.3 |
| 15 | 60-64 | M | - | - | + | - | + | - | C | Y ($> 12$) | 1.7 | 100 | 0.41 | 0.94 | 26 | 42 |
| 16 | 75-80 | P | + | + | + | - | + | - | O ($> 12$) | Y ($> 12$) | 0.29 | 547 | 0.096 | 0 | 22 | 19 |

### Antigen Specific Antibody Response

RBD-specific antibodies were measured using a quantitative ELISA assay. Twenty-five percent (4 of 16) of participants had measurable antibodies following 2 doses of mRNA vaccine (Subjects 3, 11, 12 and 16). Post-vaccine measurements were obtained 24-57 days following second dose of vaccine (one subject was drawn 103 days post vaccine). Figure 1 shows the magnitude of RBD-specific antibody response, stratified by treatment group. One of the 4 responders was treatment-naïve (Ab titer 2,981), 2 responders were under observation following previous active treatment with a geometric mean titer (GMT) of 13,068, one subject was vaccinated at greater than 12 months after treatment and the others were vaccinated between 6-12 months after treatment, and the subject with the lowest RBD-specific antibody response was on active treatment (Ab titer 331). The responder on active treatment was on the BCL-2 inhibitor, venetoclax, and had received ant-CD20 mAb treatment with rituximab 9 months prior to vaccination. Vaccinated age- and gender-matched healthy control subjects were included to establish the range of normal immune responses to mRNA COVID-19 vaccination (GMT 14,828). Baseline IgG levels were available for 3 of the 4 participants with an RBD-specific antibody response (Table 2). All had levels above 500 mg/dL (range 547-780).

**Figure 1.**
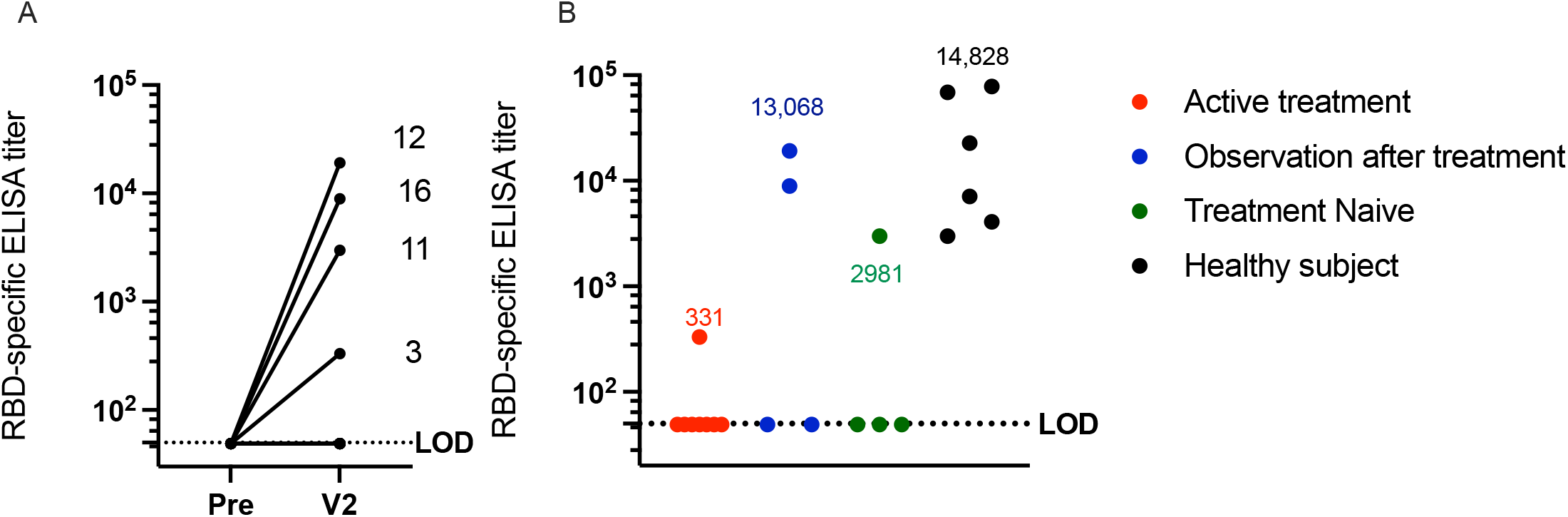
RBD-specific endpoint ELISA titer following COVID-19 mRNA vaccination. A) Pre bleed prior to vaccination and V2 following 2-dose vaccination series (24-103 days). B) RBD-specific ELISA titer stratified by treatment group, geometric mean titer (GMT) of responders shown above graph. The limit of detection, LOD, is set at 50, samples below the LOD were given an arbitrary value of 49. Healthy subject samples were taken (13-28 days) following 2-dose vaccination series.

### Antigen Specific Memory B-cell Frequency

RBD-specific memory B-cell frequencies were determined by limiting dilution assay, in which peripheral blood mononuclear cells (PBMCs) are stimulated in culture wherein memory B-cells become antibody secreting cells. The secreted antibodies can be functionally assessed for antigen-specificity by ELISA. Figure 2 shows RBD-specific memory B-cell frequency stratified by treatment group. Only one subject (Subject 12) had a detectable RBD-specific memory B-cell response of 7.49 RBD-specific MBCs per million CD19+ B-cells following a two dose COVID-19 mRNA vaccination series. This subject was 6-12 months post active treatment. Vaccinated age- and gender-matched healthy control subjects were included to establish the range of normal immune responses to mRNA COVID-19 vaccination, geometric mean frequency 4.74 (0.98 – 7.95), of note, these samples were 247-264 days following two-dose mRNA vaccination series.

**Figure 2.**
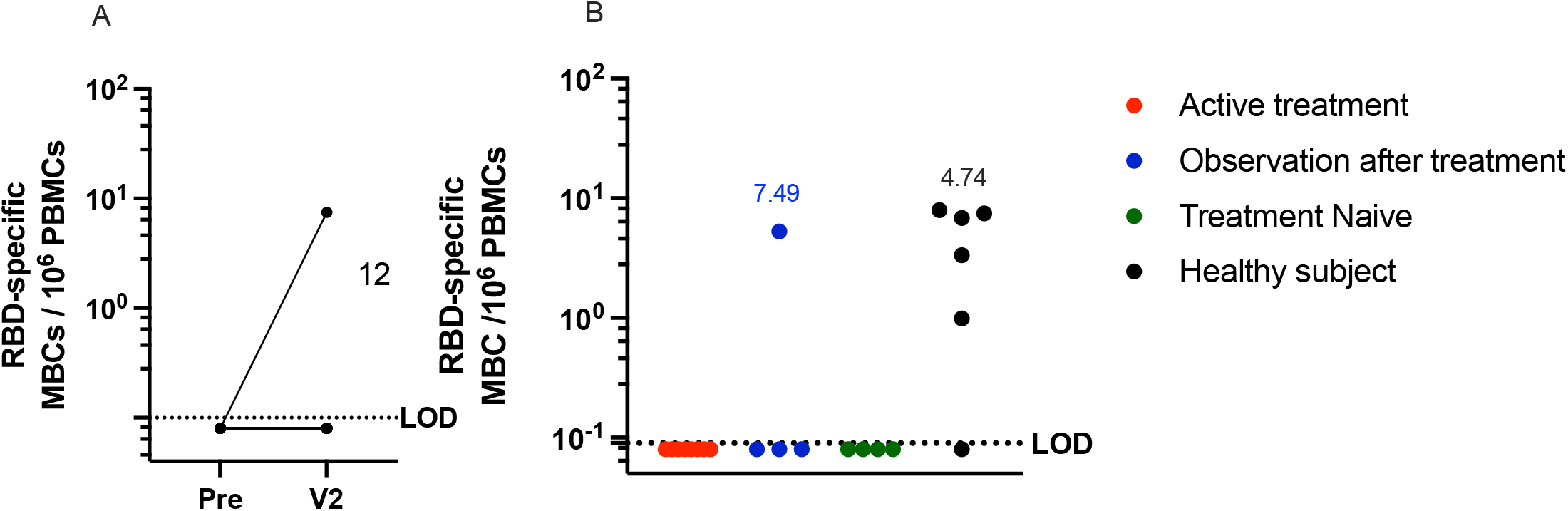
RBD-specific memory B-cell frequency per 10^6^ PBMCs following COVID-19 mRNA vaccination. A) Pre bleed prior to vaccination and V2 (24-103) days following 2-dose vaccination series. B) RBD-specific MBC frequency stratified by treatment group, geometric mean titer of responders shown above graph. Healthy subject samples were age/gender matched to CLL cohort, (247-264) post 2-dose vaccine series. Limit of detection, LOD = 0.1, an arbitrary number 0.08 was assigned to samples below the limit of detection.

### Cellular Immune Response

SARS-CoV-2 spike (S)-specific CD4+ and CD8+ T cell responses were determined by intracellular cytokine staining for IFN*γ* and TNF*α* following stimulation with overlapping peptides spanning the full-length S protein. The frequency of virus specific CD4+ and CD8 T-cells per million are shown in Figure 3 and Supplemental Table 1, both pre-vaccination and following second dose. We found that 62.5% of subjects (10/16) had CD4+ S specific responses to the vaccine, and 56% of subjects (9/16) had CD8 S specific vaccine responses. Figure 3B shows the change in S specific CD4+ and CD8+ response to vaccination, stratified by treatment group. The most subjects with a vaccine-specific CD4+ response, as well as the highest change in vaccine response was seen in the observation after treatment group. The three subjects (all in disease remission) with the most robust response have a mean expansion of 541 CD4+ T-cells/10^6^ T-cells (435-701), while the 4^th^ subject (not in remission) has a CD4+ T-cell response of only 23 CD4+ T-cells/10^6^ T-cell. The greatest change in CD8 T-cell responses was seen in the observation after treatment group (31-856) with the lowest response recorded in the treatment naïve group, where the geometric mean change in response was 132 (75-235) CD8+ T-cells/10^6^ T-cell.

**Figure 3.**
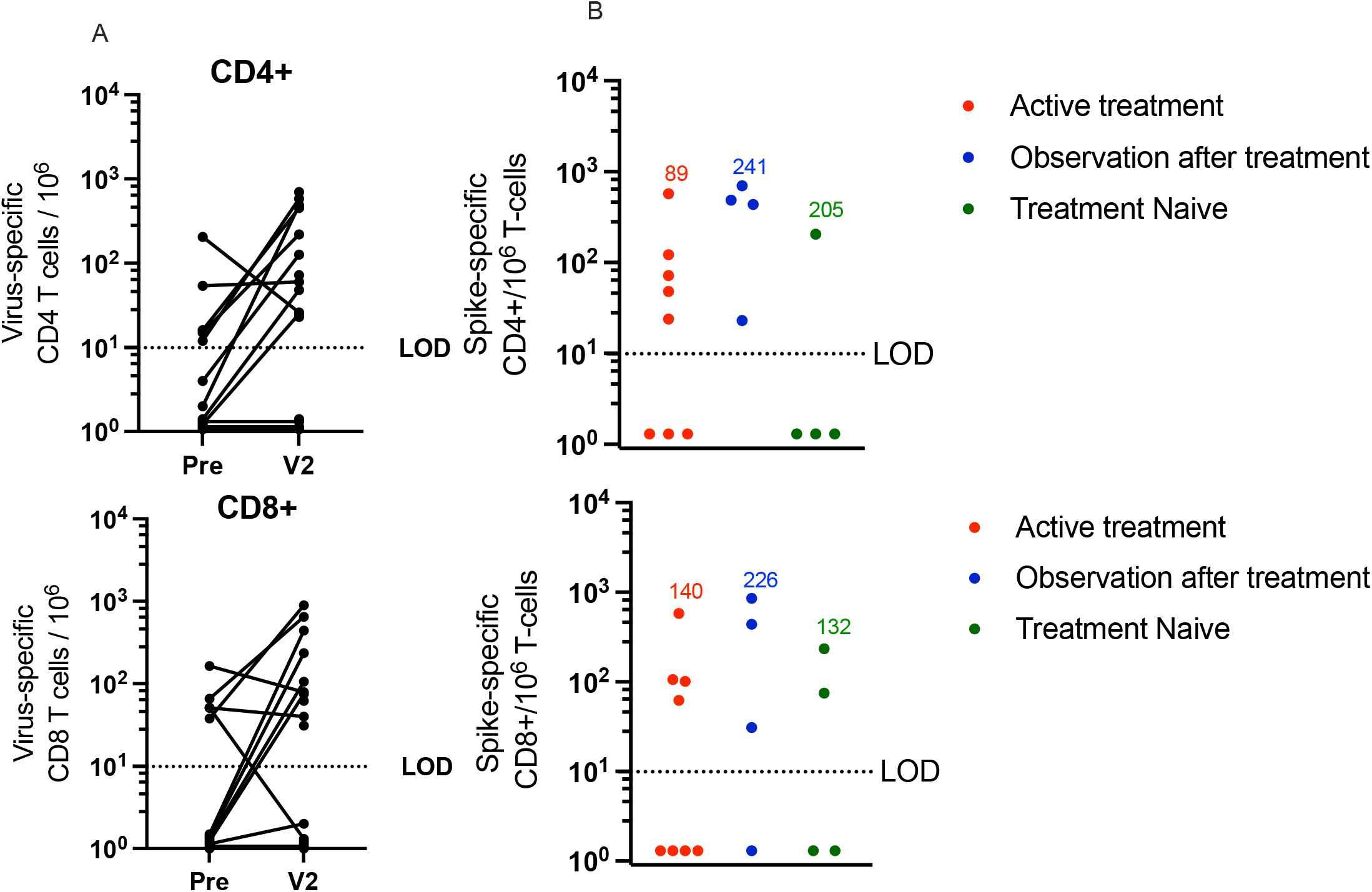
Spike-specific CD4 and CD8 T-cell frequency per 10^6^ T-cells following COVID-19 mRNA vaccination. A) Pre bleed prior to vaccination and V2 following 2-dose vaccination series (24-103 days). B) Spike-specific CD4+ and CD8+ response to vaccination, the increase in T-cell expansion from baseline, stratified by treatment group, geometric mean of responders shown above graph. Limit of detection (LOD =10), For subjects without vaccine specific response, a arbitrary value between 1.1-1.5 was assigned.

### Recall humoral immune response

In order to better characterize overall humoral immunity in our study subjects, we also evaluated their antibody-mediated recall response to measles virus, a virus antigen that most people are exposed to in childhood either through vaccination or natural infection. Both serum (LLPC-derived) antibodies and measles-specific memory B-cells (Figure 4) were examined. Detectable measles-specific antibodies at or above the limit of detection (1:100) was observed in 81% (13 of 16) of subjects with the highest measles specific antibody titers observed in the observation after treatment and the age-gender matched healthy control subjects (GMT 3670 and 3432). While measles-specific memory B-cells were only observed in 25% (4 of 16) subjects, two currently on active treatment with BTKi (geometric mean frequency of responders 1.31) and two treatment-naïve (geometric mean frequency of responders 2.4), and 3 of the 4 subjects who responded had never received CD20 mAb treatment. However, these frequencies are much lower than that observed in age-gender matched healthy control subjects, geometric mean frequency 78.4 (17.3 – 451.5).

**Figure 4.**
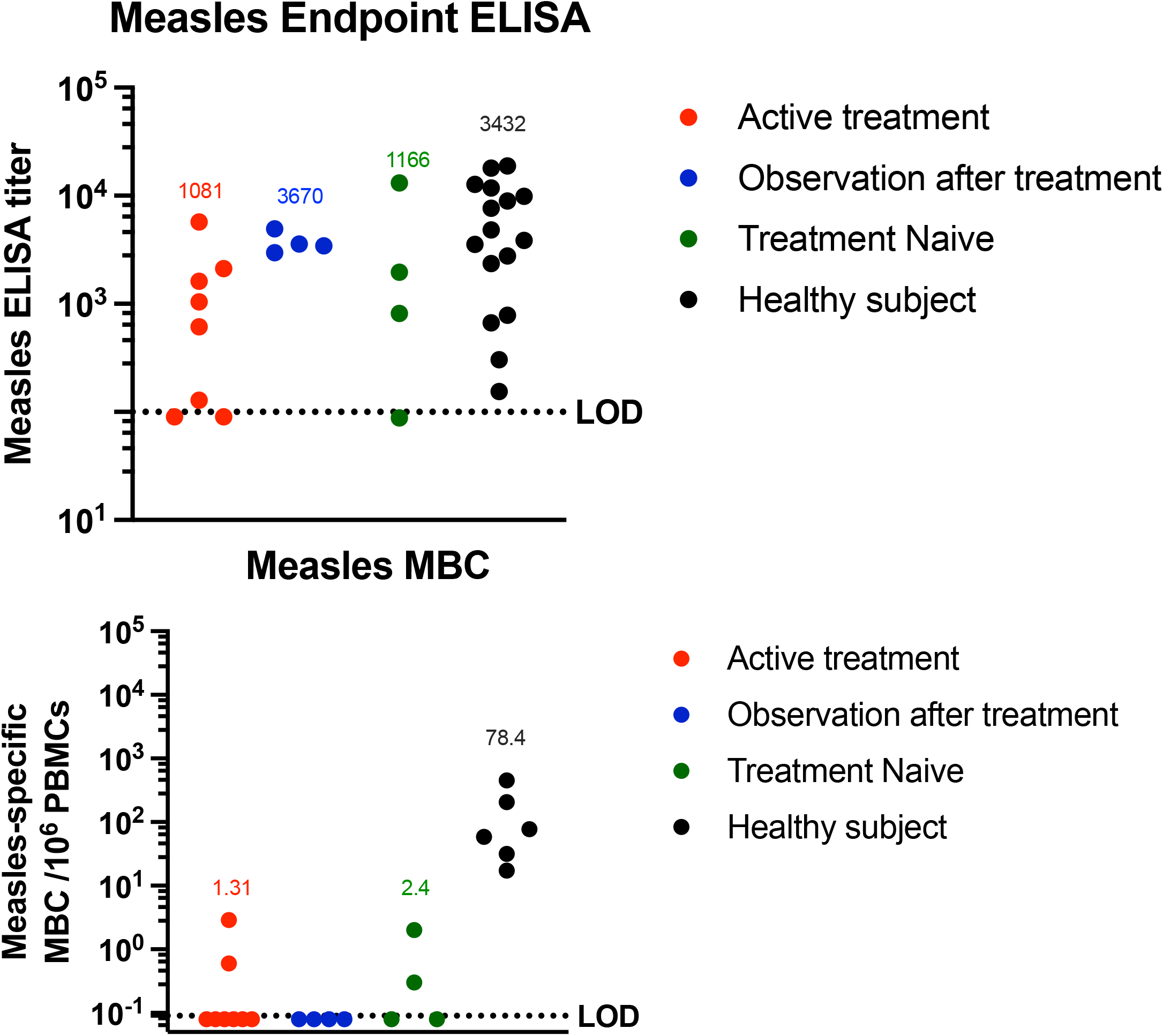
Humoral immune recall response to a childhood antigen, measles, in CLL subjects and age-gender matched healthy controls. A) Measles-specific endpoint ELISA titer stratified by treatment group. Limit of detection (LOD = 100) Samples below the limit of detection assigned an arbitrary value of 80. Geometric mean titer (GMT) of responders shown above the graph for each group. B) Measles-specific MBC frequency stratified by treatment group, geometric mean frequency of responders shown above graph. Limit of detection, LOD = 0.1 an arbitrary number between .05 and .1 was assigned to those samples.

### Descriptive statistical analysis to understand relationship between clinical factors and immune response

We investigated the relationship between baseline laboratory values and humoral or cellular immune response to vaccination as part of a pre-specified analysis plan. We compared mean percentage of CD19+ B-cells, IGD+27-naïve B-cells, IGD+27+ non-switched B-cells, IGD-27+ switched memory B cells and CD5+CD19+ B1 B-cells between responders and non-responders for antibody response, CD4+ response, CD8 response and memory B-cell response respectively. There were no statistically significant differences between any of the compared groups (Supplementary Table 2 and 3). However, responders had lower mean values for CD19+ B-cells (23.3% vs. 34.4%), CD5+19+ B-cells (19.5% vs. 32.8%), and IGD-27+ switched memory B-cells (5.3% vs. 8.4%) and higher mean values for IGD+27-naïve B-cells (29.9% vs. 24.6%) and IGD+27+ non-switched B-cells (45.3% vs. 40.8%).

Mean values of baseline IgG level and absolute lymphocyte count did not show statistically significant differences between responders and non-responders. (Supplementary Table 3). Participants with antibody response had a median IgG level of 593 (range 547-780) compared to 333 (range 85-918) in participants without antibody responses. There was also no statistically significant difference regarding previous anti-CD20 mAb treatment, P value 0.3 – 1.0 depending on antibody, T-cell or MBC immune response (Supplementary Table 3). One participant (subject 15) subsequently developed symptomatic SARS-CoV-2 infection that was confirmed by polymerase chain reaction. This patient was on active treatment with BTKi and prior to infection, had not developed a humoral immune response or CD4+ specific response, however they did generate a CD8+ specific response of 101 CD8+ T-cells/10^6^ T-cells after vaccination.

## Discussion

The results presented in this study indicate that the cellular and humoral immune responses to COVID-19 vaccination is impaired in CLL subjects. This is consistent with previous studies^18,19,20^ which observed a seroconversion rate from 23.1% to 50% overall in CLL subjects following mRNA vaccination, further reduced when stratified by treatment status. Here, we observe a 25% seroconversion rate as measured by quantitative ELISA. The 4 patients with vaccine-mediated antibody responses were diverse with 1 treatment naïve patient, 1 patient on current treatment with bcl-2 inhibitor and 2 patients under observation. For patients under observation, 1 patient was in remission while the other patient had relapsed disease. When stratified by treatment status, 50% of the subjects currently on observation following treatment seroconverted compared to 12.5% of subjects currently on active treatment. Of the responders (4/16) one had never received anti-CD20 mAb treatment and three had received it over 12 months prior, consistent with previous studies.^21^ In an attempt to identify potential predictors of vaccine response, we evaluated a number of clinical factors as well as a broader range of immune profiling. Although no significant differences were appreciated, the responders had overall higher IgG serum levels, and lower ALC, mean B-cell percentage, class switched MBCs, and B1-B-cells when compared to non-responders (Supplementary Table 2 and 3). A larger sample size is needed to determine if any of these clinical parameters can serve as predicters of antibody response to novel antigen vaccination.

Interestingly, only one subject exhibited an RBD-specific memory B-cell response (Subject 12). This subject is in observation after treatment, currently in disease remission, with bcl-2 inhibitor treatment occurring > 6 months prior to vaccination in combination with an anti-CD20 mAb treatment given > 12 months prior to vaccination. This subject had not been treated with a BTKi. Baseline (prior to vaccination) evaluation of lymphocyte populations revealed normal ranges for CD19+ B-cells (8%), as well as CD4+ (42%) and CD8+ (15%) T-cells. A recent study found that CD19+ B-cells were correlated with SARS-CoV-2 specific antibody response.^22^ The observation that 3 of 4 patients with an RBD-specific antibody response did not have detectable RBD-specific memory B-cells is notable and suggests that additional studies evaluating antibody persistence as well as memory B-cell response are needed.

We hypothesize two reasons for this lack of memory B-cell response. First, it is possible that the positive antibody titers only represent short-lived plasma cell/plasmablast antibody response without persistence of long-lived antigen-specific memory cells or long-lived plasma cells. Short-lived plasma cells soon after infection or vaccination do not persist and aren’t thought to require the formation germinal centers or require CD4+ T-cell help.^23^ Of note, all subjects who had an RBD-specific antibody response also had a spike-specific CD4+ T-cell response, indicating that a population of T-helper cells was available for B-cell priming. If what we are seeing are in fact antibodies secreted by short-lived plasma cells, they are not committed to memory and end up undergoing apoptosis, although the antibodies themselves would persist in the serum, IgG-specific antibodies have a half-life of about 26 days.^24^ The other possible explanation is that germinal center formation is taking place, but that the process is inherently slow in CLL individuals with impaired humoral immune responses. Both of these hypotheses could be tested with longitudinal sampling from this cohort of CLL subjects.

SARS-CoV-2 Spike reactive T-cells were present at baseline in some of the subjects. Specifically, Subjects 5 and 8, which exhibited no expansion of spike-responsive CD4+ T-cells following vaccination and Subjects 3 and 13 which exhibited no boost in spike-reactive CD8+ T-cells following vaccination. This is consistent with previous reports of spike reactive T-cells in naïve individuals without prior antigen exposure.^25^

Overall, the cellular immune response appeared to be much less impaired, even fairly robust in these CLL subjects compared to humoral immune response. Robust functional T-cells were also observed in a large cohort study of 195 subjects with hematologic malignancies in which 53% of subjects had SARS-CoV-2 specific, IFN*γ* secreting T-cells.^22^ In the study presented here, we observed 62.5% (10/16) of subjects had a vaccine S-specific CD4+ response and 56% (9/16) had a CD8+ T-cell response. Of these subjects, 4 had a S-specific CD4+ response alone, 3 had a S-specific CD8+ response alone, and 6 subjects had both a CD4+ and CD8+ response. None of the subjects with a CD8+-only response had a humoral immune response to vaccination. Four of the 10 CD4+ responders seroconverted, providing supporting evidence for the importance of CD4+ T-cell help in generating a B-cell response. The robust cellular responses seen post vaccine are consistent with results seen after natural infection in CLL patients with a recent study observing that 82% of subjects developed S-specific T cell responses.^26^ However, unlike these results, the same study saw a high seroconversion rate in naturally infected CLL subjects (82%) by semi-quantitative antibody test. Interestingly, some subjects were negative for S-antibodies at early timepoints, but seroconverted later, or were negative for serum antibodies, but positive for antibodies in the saliva (IgA), a population of antibodies not explored in this investigation. Overall these findings suggest that natural infection may lead to a more robust immune response in subjects with CLL compared to vaccine response ^26^ Important to note, the current study focused on elucidating IgG-specific antibodies, both in the serum and MBC-derived.

Like the humoral response, the cellular response appears to be highly treatment dependent. The most responders as well as the highest response was seen in the observation after treatment group, for both CD4+ and CD8+ T-cells. The lowest CD4+ response was seen in subjects undergoing active treatment and treatment naïve for the CD8+ T-cell response. SARS-CoV-2-specific CD8+ T-cell responses and CD8+ T-cell count was associated with improved survival in patients with hematologic malignancies hospitalized with SARS-CoV-2 infection.^27^ This included patients on CD20 depleting therapy and patients that had received monoclonal antibody with impaired humoral response. Data in healthy participants receiving SARS-CoV-2 vaccine suggest rapid CD4+ T-cell response after single vaccine however CD8+ T-cell response only after second dose.^28^ In CLL patients with impaired humoral response, it is unknown whether CD8+ response in CLL patients demonstrate long term effector T-cell protection, and studies on whether additional booster vaccine induce such protection are needed.

Active treatment with BTK inhibitors like Ibrutinib has a significant impact on B-cell survival, differentiation and the development of an antigen-specific antibody response to novel antigen exposure either by natural infection or vaccination. B-cells are dependent on BTK signaling for differentiation and proliferation signals, therefore subjects on BTK inhibitors experience greater immune dysfunction. Immune response to novel antigens, either by natural infection or vaccination is severely limited in these subjects,^15^ however recall to previously encountered antigens such as those seen during childhood appear to remain largely intact. Five of the 7 participants (71%) on BTKi had cellular immune response with CD4+ and/or CD8+ T-cells. This suggests that cellular responses may be preserved even in CLL patients on BTKi. Whether this finding translates to an effective T-cell response that provides protection against infection even in the absence of humoral response is of clinical interest. The impact of prolonged treatment vs. shorter-term BTK inhibition on immune responses is unknown. However, clinical data suggests some improvement in humoral immunity with prolonged treatment (> 6 months), including a decrease in the number of infections and an increase in serum IgA levels.^29^ The immune disruption experienced by CLL subjects is not limited to the humoral immune system, T-cells can also be disrupted in CLL subjects and even further affected by BTK inhibitors^30,31^ although that was not observed in this study. Given BTK inhibitors are administered daily on a continuous basis, further studies that evaluate timing of vaccines, as well as interruption of ongoing BTKi therapy in an attempt to enhance vaccine response are warranted. This is a paradigm that has shown some success in patients with rheumatologic disease on treatment with immunosuppressive therapies.^32^ Bcl-2 is a protein regulator of apoptosis and preclinical data suggests bcl-2 inhibition affects T-cell function.^33^ The impact of ongoing bcl-2 inhibition with venetoclax as a single agent remains an unanswered question worthy of additional study.

A recent study^15^ reports impaired vaccine response to novel antigens in CLL patients. Vaccination with recombinant hepatitis B vaccine (HepB-CpG) led to a response in 28.1% of treatment naïve subjects and only 3.8% of patients on BTK inhibitors. When this was compared to humoral response to known antigens represented by recombinant zoster vaccine (RZV) the response was 41.5% in subjects on BTK inhibitors and 59.1% for treatment naïve subjects indicating that BTK inhibitors disrupt the generation of a novel immune response, but don’t necessarily significantly interfere with recall response. In this investigation, we explored the recall response to measles virus, a common childhood antigen exposed to either through vaccination or natural infection in subjects born before 1963.^34^ Here, we observed that 81% of our subjects were seropositive for measles serum antibodies, with all but three subjects exhibiting a response above background (limit of detection). Of those, two subjects (Subject 3 and 10) are currently on active treatment with bcl-2 inhibitor and BTKi respectively and one subject is treatment naïve. This is a slightly higher response rate than was observed in a recent cross-sectional study of 959 patients^34^ which detected an overall 75% measles seropositivity rate in cancer subjects and lower (63% seropositivity) in those with hematological malignancies. Overall, the antibody response to measles seems largely unaffected in CLL subjects, indicating that long-lived plasma cells responsible for maintaining circulating serum antibodies remain stable throughout CLL immune dysfunction and treatment. On the other hand, the memory B-cell recall response to measles was highly disrupted in these subjects. Only 25% retained a detectable population of measles-specific memory B-cells, of these four subjects, two were on active treatment (Subject 5 and 6) and two were treatment naïve. Even though a population of measles specific MBCs was detected in these subjects, the frequencies were much lower than those observed in age/gender matched healthy subject controls (geometric mean frequency 78.4). The subjects’ measles specific B-cell responses were not unexpected as CD20 mAb treatment which eleven of our subjects received, depletes circulating B-cells.. These findings may have clinical implications especially in the setting of vaccines that are not associated with durable antibodies where the combination of memory b-cell depletion and diminishing antibody response may abrogate viral-specific humoral immunity.

Study limitations include a small sample size and lack of long-term follow up to identify long-lived humoral response. None of the 16 subjects enrolled in this study tested positive for SARS-CoV-2 prior to enrollment in the study/vaccination, however, although testing for prior infection via evaluation of RBD antibodies in the serum was performed prior to vaccination in 12 of the 16 subjects, evaluation and anti-nucleocapsid antibodies which is more specific for prior infection was not performed in any subjects. Long-term clinical outcomes to evaluate vaccine efficacy in relation to vaccine response is also needed.

In conclusion, the results of this study provide a thorough evaluation of the humoral and cellular immune response to initial 2-dose mRNA COVID-19 vaccine series in CLL patients. Our results highlight the limitations of serology studies alone in defining vaccine-mediated immune responses, particularly in this immune-dysregulated patient population. Larger longitudinal studies which incorporate clinical outcomes in vaccinated CLL patients as well as study the impact of a third, booster or heterologous vaccine are needed.

## Methods

### Study Design

This study was conducted at a single site at Oregon Health and Science University. The protocol was approved by the Oregon Health and Science University Institutional Review Board (OHSU IRB# 21230). All participants gave written informed consent before enrolling in the study. This vaccine arm of the cohort is part of a larger biobank study on human immune response during and following acute viral infection and vaccination against novel viruses. SARS-CoV-2 vaccines were administered through standard procedures as part of the Oregon’s COVID-19 Vaccination Plan and not as part of the study. On initial enrollment, demographics, CLL disease characteristics, and treatment details were collected (Table 1), and baseline laboratory values were obtained (Table 2), including SARS-CoV-2 spike antibody titer, serum IgG, a complete blood count, and multicolor flow cytometry measuring immune cell populations (Table 2). One-month following completion of 2-dose mRNA vaccination series, follow up draws were completed and vaccine-specific immune response was evaluated. Study protocol includes additional serology studies at 3 months, 6 months, and 1 year following vaccine.

### Participants

Eligible participants were adults 18 years or older with confirmed diagnosis of CLL or small lymphocytic lymphoma (SLL) who did not have a history of previous SARS-CoV-2 infection or vaccination. Subjects were excluded if they had received anti-CD20 therapy within 6 months of enrollment. Prior intravenous immune globulin (IVIG) therapy was not an exclusion criterion.

### Healthy subject controls

Age and gender matched controls who received a two-dose mRNA (Pfizer-BioNTech) vaccine in January – February 2021were enrolled in a vaccine study approved by the Oregon Health and Science University Institutional Review Board. Serum from twelve age/gender matched controls (13 – 28 days following 2^nd^ dose) and PBMCs (247 – 267 days following 2^nd^ dose) from six age/gender matched controls were used for determining RBD and measles specific antibody responses in immunocompetent subjects.

### Statistical Analysis

Mean and median lymphocyte panel laboratory findings were determined for each positive and negative antibody response, cellular immune response, and MBC frequency. Group means were compared using an independent t-test, except where applicable and noted, the Satterthwaite test statistic was used instead. Mean, median, and independent group t-tests were also calculated for IgG and ALC values collected at the time of vaccination. The relationship between prior CD20 mAB treatment and current treatment status with antibody, MBC frequency, and cellular immune responses were similarly assessed using a chi-square test, or Fisher’s exact, where applicable. All statistical analyses were performed in SAS version 9.4.

### Sample processing

After obtaining informed consent, at the time points indicated, 10mL of whole blood was collected for serum (BD Vacutainer® Red Top Serum Tubes) and 40mL of whole blood was collected for PBMCs and plasma (BD Vacutainer® Lavender Top EDTA Tubes). Plasma and serum samples were centrifuged for 10 minutes at 1000 x G, heat inactivated and stored at -20C. PBMCs were isolated and stored in liquid nitrogen until needed.

### Antigen-specific plasma endpoint ELISA

ELISAs were performed as previously described^35^, Briefly, ninety-six well ELISA plates (3590, Corning) were coated with 100 μL recombinant RBD protein (Provided by Dr. David Johnson) at a concentration of 0.5 μL/mL or 50 μL of measles virus antigen (MyBioSource, MBS239121) at a concentration of 4 μg/mL prepared in PBS and the plates were incubated overnight at 4°C. Coating antigen was removed, plates were washed once with PBS-T containing 0.05% Tween (wash buffer) and blocked for 1 hour at RT with 5% milk prepared in PBS-T containing 0.05% Tween (dilution buffer).

Plasma in dilution buffer, 100 μL of 1:50 (RBD) or 1:30 (measles) dilution was added to each well. Plasma samples were serially 3-fold diluted in dilution buffer. Plates were incubated at RT for 1 hour. The plates were washed 3 times with wash buffer and 100 μL of 1:3000 dilution of anti-human IgG (H+L) HRP (Novus, NBP1-73319) detection antibody was added and incubated at RT for 1 hour. After rinsing (3X) with wash buffer, 100 μL of colorimetric detection reagent containing 0.4 μg/ml o-phenylenediamine and 0.01% hydrogen peroxide in 0.05 M citrate buffer (pH 5) were added and the reaction was stopped after 20 minutes by the addition of 100 μL 1 M HCl. Optical density (OD) at 492 nm was measured using a CLARIOstar ELISA plate reader. Antibody titers were determined by logarithmic transformation of the linear portion of the curve with an endpoint of 0.1 optical density units.

### MBC limiting dilution analysis

PBMCs were thawed and resuspended in LDA media, RPMI 1640 (Gibco), 1×Anti-Anti (Corning), 1X non-essential amino acids (HyClone), 20 mM HEPES (Thermo Scientific), 50μMβ-ME, and 10% heat-inactivated FBS (VWR). Cells were cultured in serial 2-fold diluted doses (10 wells per dose), starting with 3-5 × 10^5^ PBMCs per well at the highest dose. In 96-well round-bottom plates in a final volume of 200μl per well^36^. Cells were incubated with IL-2 (Prospec) 1000U/ml and R848 (InvivoGen) 2.5ug/m.^37^ To determine background absorbance values, supernatants were used from 8 wells unstimulated PBMCs only. Plates were incubated at 37 °C and 5% CO^2^ for 7 days. Stimulation was determined by running total IgG ELISAs. Antigen-specific MBC frequencies were calculated by assaying LDA supernatants by antigen-specific ELISAs. MBC precursor frequencies were calculated by the semi-logarithmic plot of the percent of negative cultures versus the cell dose per culture, as previously described ^38^ and frequencies were calculated as the reciprocal of the cell dilution at which 37% of the cultures were negative for antigen-specific IgG production. Cell doses which yielded 0% negative wells were excluded, since this typically resides outside of the linear range of the curve and artificially reduced the MBC precursor frequency. For subjects with low frequency of antigen-specific antibody secreting cells frequency was determined by number of positive wells divided by the total number of IgG positive secreting wells, multiplied by one million, giving a frequency per million PBMCs stimulated.

### Antigen-specific ELISA for LDA

Ninety-six well half-well ELISA plates (Greiner Bio-one) were coated with 50uL of 0.5 ug/mL antigen in PBS, recombinant RBD protein (provided by Dr. David Johnson) and measles virus antigen (MyBioSource, MBS239121). Plates were incubated overnight at 4°C, washed once with PBS-T containing 0.05% Tween (wash buffer) and blocked for 1 hour with 5% milk prepared in PBS-T containing 0.05% Tween (dilution buffer). 20 uL of LDA supernatants were added to each well. Plates were incubated at room temperature (RT) for 1 hour, washed 4 times with wash buffer, and 50 uL of 1:3000 dilution of anti-human IgG-HRP (BD Pharmingen, 555788) detection antibody was added and incubated at RT for 1 hour. Plates were washed 4 times with wash buffer, 50 uL of colorimetric detection reagent containing 0.4 mg/ml o-phenylenediamine and 0.01% hydrogen peroxide in 0.05 M citrate buffer (pH 5) were added and the reaction was stopped after 20 minutes by the addition of 50 uL 1 M HCl. Optical density (OD) at 492 nm was measured using a CLARIOstar ELISA plate reader. Positive wells were determined as wells 2-fold above background (unstimulated PBMC wells).

### Spike-specific simulation and intracellular cytokine staining

PBMCs were thawed at 37°C, washed and resuspended in RPMI1640 (Corning) supplemented with 5% FBS (Hyclone), Glutamine (Corning), HEPES (Lonza) and Pen Strep (Gibco). Cells were stimulated at 1 million cells/well in 200 μl in 96 well round bottom plates (Corning) at 37°C/6% CO_2_ with 2 peptide pools (peptides 1-90 and 91-181 at 0.5 μg/ml of each peptide) of overlapping (10AA) 17mers representing the SARS-CoV-2 Spike protein (BEI Resources). All conditions contain a final DMSO concentration of 0.045%. Positive control wells were stimulated with 0.04 μg/ml anti-CD3 (HIT3a NA/LE, BD Biosciences) in media containing 0.045% DMSO or incubated with 0.045% DMSO media alone to assess spontaneous production of cytokines. After 1 hour, Brefeldin A (Sigma) was added to a final concentration of 2 μg/ml and stimulations were continued for 5 hours. Intracellular cytokine staining was performed as described previously.^39,40^ After stimulation, cells were stained overnight at 4°C with anti-CD8, anti-CD4+ (2ST8.5H10 and L200 BD Biosciences) and Aqua live/dead stain (InVitrogen) in PBS + 1% FBS + 0.1 mg/ml MsIgG. Following overnight incubation, cells were fixed with 2% formaldehyde in PBS. After staining with anti-IFN*γ* and anti-TNF*α* (4S.B3 and Mab11 from eBioscience)) in PermWash cells were washed with PermWash, PBS + 1% FBS and fixed with 2% formaldehyde in PBS. Data was acquired on an LSR Fortessa (Becton Dickenson) and analyzed using FlowJo software (Becton Dickenson). Cytokine expression in medium +DMSO alone cultures was subtracted from peptide-stimulated cultures to calculate peptide-specific cytokine expression. Responses to both peptide pools were added together to yield the total frequency of SARS-CoV-2-specific cytokine producing CD4+ and CD8+ T cells.

## Data Availability

All data produced in the present work are contained in the manuscript

## Acknowledgements

The authors would like to thank the subjects for participating in this research study. This work was funded in part by National Institute of Allergy and Infectious Diseases NIAID 1R01AI145835 (WBM), by US National Institute of Health grant P51 OD011092 (MKS), and endowed funds from the Knight Cancer Institute’s Scholar Award for Leukemia and Lymphoma Research (SES). The funders had no involvement in study design; in the collection, analysis and interpretation of data; in the writing of the report; or in the decision to submit the article for publication.

## Author Contributions

**Z.L. Lyski**: Formal analysis, Investigation, Visualization, Writing-original draft, editing and reviewing. **M.S. Kim**: Investigation, Project administration, Resources, Visualization, Writing-original draft. **D. Xthona Lee**: Investigation, formal analysis **H-P. Raué**: Formal analysis, Investigation, Writing-review & editing. **V. Raghunathan**: Investigation, Project administration, recourses. **J. Griffin** Project administration, Resources. **D. Ryan**: Project administration, Resources. **A.E. Brunton**: Formal analysis, Resources. **M.E. Curlin**: resources, Writing – review & editing **M.K. Slifka**: Funding acquisition, Supervision, Writing – review & editing **W.B. Messer**: Conceptualization, funding acquisition, Supervision, Writing – review & editing **S.E. Spurgeon**: Conceptualization, funding acquisition, Project administration, supervision, Writing – original draft, writing – review & editing.

## Figure Legends and Tables

**Supplementary Table 1**. T-cell specific response per subject, pre and post vaccine. CD4 and CD8 spike-specific T-cell response per subject per 10^6^ T-cells. N: Treatment Naïve, C: Currently on treatment, O (6): Observation, last treatment within 6 months, O (6-12): Observation, 6-12 months since last treatment, O (>12): Observation, more than 12 months since last treatment.

**Supplementary Table 2**. Specific B-cell populations (%) stratified by antibody, CD4, CD8, and MBC responders and non-responders.

**Supplementary Table 3**. Clinical markers, prior CD20, current treatment group, serum IgG at baseline as well as ALC at baseline, stratified by antibody, CD4, CD8, and MBC responders and non-responders.

