## Supplementary figures and images for "Cellular and humoral Immune response to mRNA COVID-19 vaccination in subjects with chronic lymphocytic leukemia"

### Supplemental table 1

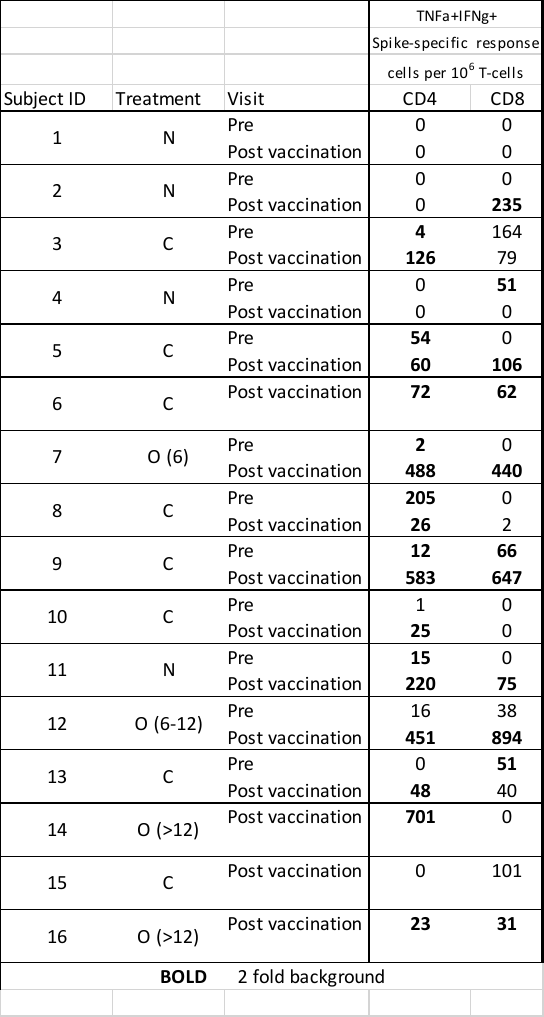

### Supplemental table 2

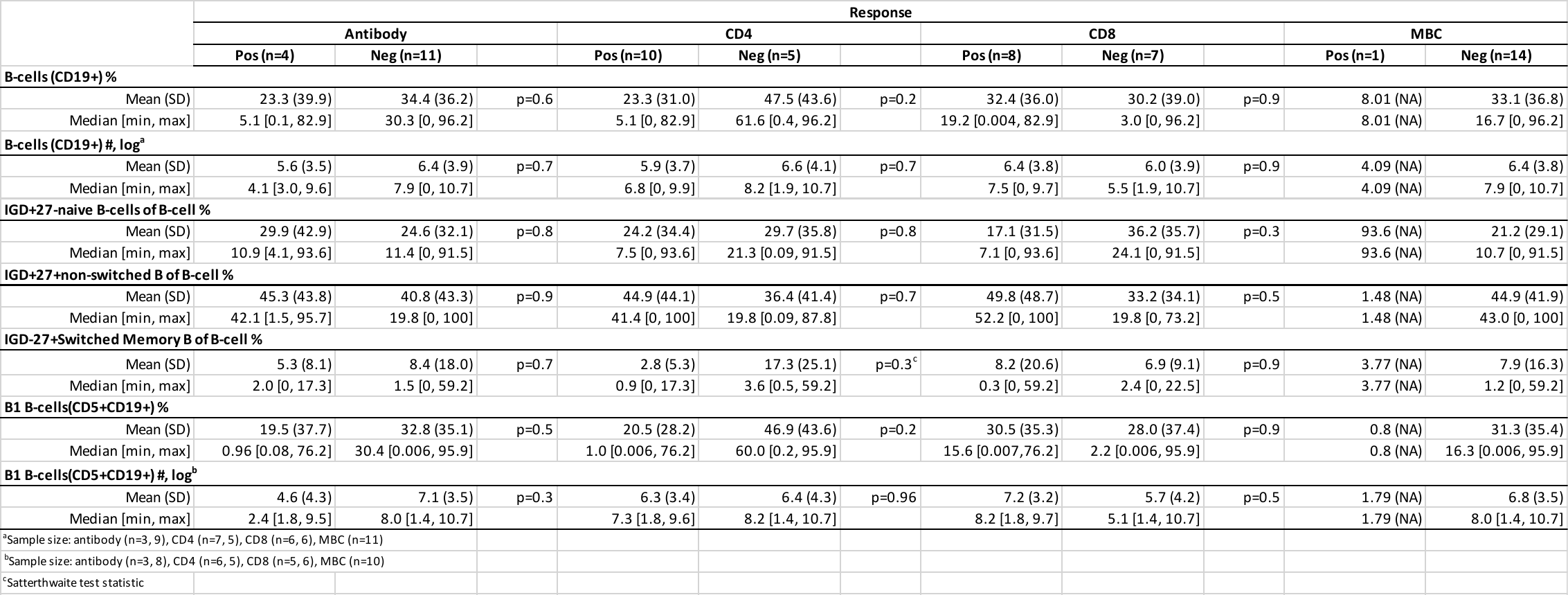

### Supplemental table 3

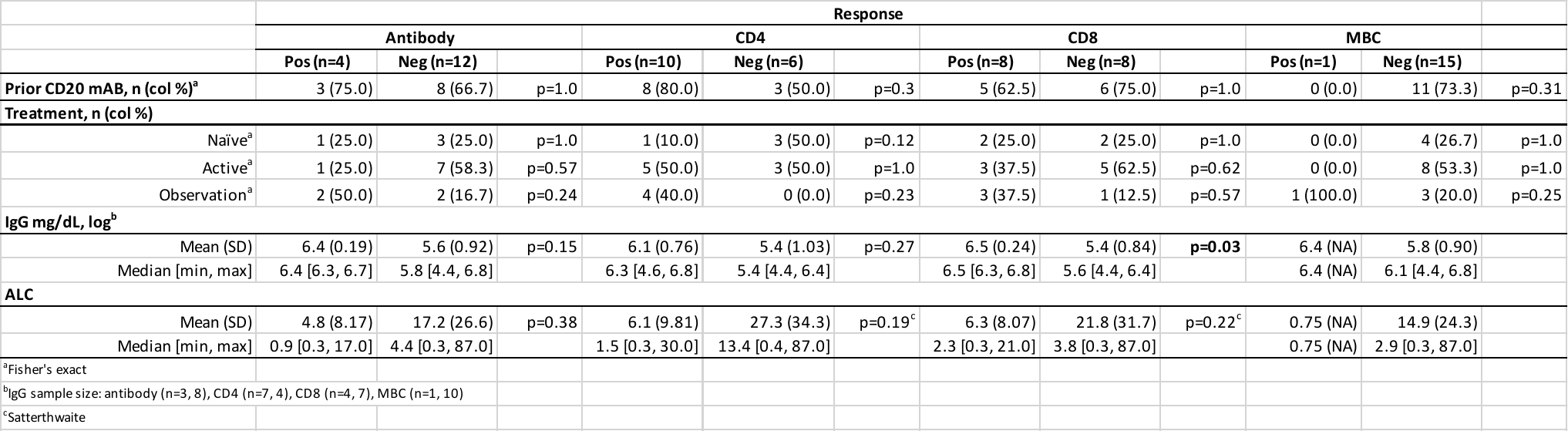
